# Hypertensive disorders of pregnancy and risk of cardiovascular disease: A Mendelian Randomisation study in the UK Biobank

**DOI:** 10.1101/2023.03.14.23287281

**Authors:** Lena Tschiderer, Yvonne T van der Schouw, Stephen Burgess, Kitty WM Bloemenkamp, Lisa Seekircher, Peter Willeit, N Charlotte Onland-Moret, Sanne AE Peters

**Author notes:** **Correspondence to:** Sanne AE Peters, Julius Centre for Health Sciences and Primary Care, University Medical Centre Utrecht, Utrecht, the Netherlands.

## Abstract

**Background:** Observational studies show that hypertensive disorders of pregnancy (HDPs) are related to unfavourable maternal cardiovascular disease (CVD) risk profiles later in life. We investigated whether genetic liability to pre-eclampsia/eclampsia and gestational hypertension is associated with CVD risk factors and occurrence of CVD events.

**Methods:** We obtained genetic associations with HDPs from a genome-wide association study and used individual-participant-data of women and men from the UK Biobank to obtain genetic associations with CVD risk factors and CVD events (defined as myocardial infarction or stroke). The rationale for including men and nulligravidae was to study genetic liability to HDPs and CVD risk without experiencing the underlying phenotype. We applied Mendelian Randomisation analysis using inverse variance weighted regression in our primary analysis.

**Results:** We included 264,160 women (mean age 56.4 [SD 8.0]) and 223,043 men (mean age 56.7 [8.2]) with available genetic data. Genetically proxied pre-eclampsia/eclampsia was related to higher risk of CVD with odds ratios of 1.20 (1.01, 1.43) and 1.29 (1.08, 1.53) in women and men, respectively. For genetically proxied gestational hypertension, odds ratios for CVD were 1.22 (1.10, 1.36) and 1.28 (1.16, 1.42) in women and men, respectively. Furthermore, genetically proxied HDPs were associated with higher levels of systolic and diastolic blood pressure and younger age at hypertension diagnosis in both women and men.

**Conclusions:** Genetic liability to HDPs including pre-eclampsia/eclampsia and gestational hypertension is associated with higher CVD risk, implying biological mechanisms relating to these disorders are causally related to CVD risk in both women and men.

## Introduction

Hypertensive disorders of pregnancy (HDPs) affected one in eight hospital deliveries in 2019 and came along with one in four maternal deaths between 2017 and 2019 in the US.^1^ Globally, there were 18.1 million incident cases of hypertensive pregnancy disorders in 2019.^2^ They can express as different phenotypes including gestational hypertension, pre-eclampsia/eclampsia, and HELLP syndrome. Pre-eclampsia is a multi-organ disease originating from a dysfunction of the placenta and affects approximately one in twenty deliveries.^3,4^ The International Society for the Study of Hypertension in Pregnancy defines pre-eclampsia as gestational hypertension with one or more of the following concomitants: (1) proteinuria, (2) other maternal organ dysfunction (acute kidney injury, liver involvement, neurological or haematological complications), or (3) uteroplacental dysfunction.^5^ Eclampsia is a progression of pre-eclampsia, in which women additionally experience seizures.^6^ Gestational hypertension is defined as *de novo* hypertension at ≥20 weeks of gestation without the additional signs of pre-eclampsia.^5^

A range of risk factors for HDPs have been proposed including higher pre-pregnancy body mass index (BMI), pre-pregnancy diabetes mellitus, chronic hypertension, chronic autoimmune disease, and maternal age.^7,8^ Furthermore, HDPs are related to multiple immediate, short-term, and long-term health concerns, which can affect the mother and the foetus and neonate.^9^ Short-term outcomes are, for instance, stillbirth and preterm delivery,^9^ whereas on the long term HDPs are associated with maternal cardiovascular risk factors after pregnancy including diabetes mellitus, hyperlipidaemia, and hypertension.^9^ Observational studies have also reported a relation between HDPs and higher risk of maternal cardiovascular disease (CVD) events later in life,^9^ which has been confirmed in co-sibling analyses^10^ and by a previous Mendelian Randomisation (MR) study that was restricted to sex-combined genetic associations with CVD events.^11^

However, the specific biological mechanisms behind HDPs and CVD risk are not entirely clear. As HDPs and CVD share a considerable amount of risk factors, the theory emerged that both phenotypes are expressions of the same disease pathway at different stages in life,^12^ and that pregnancy enables earlier identification of women at higher CVD risk.^13^ Apart from that, HDPs could also cause long-term vascular damage leading to a higher CVD risk later in life.^13^

To better understand the role of HDPs in the development of CVD, we conducted a MR analysis of 487,203 women and men from the UK Biobank with the aim to estimate the sex-specific relation of genetic liability to pre-eclampsia/eclampsia and gestational hypertension with CVD events, blood pressure-traits, and lipid-, liver-, and kidney-related cardiovascular risk factors. The rationale for also analysing men and nulligravidae was to study the role of genetic liability to HDPs in cardiovascular risk without experiencing the underlying phenotype, which could help understand whether biological mechanisms related to HDPs or the diseases themselves are responsible for higher CVD risk.

## Methods

This analysis adheres to the STROBE-MR statement (**Table S1**).^14^ Please see the Major Resources Table in the Supplemental Materials.

### Study design and data sources

The present study included data from the UK Biobank, of which details have been described previously.^15^ Briefly, the UK Biobank is a large-scale prospective study in the general population of the UK, in which over 500,000 individuals aged 40 to 69 years were recruited between 2006 and 2010.^15^ The UK Biobank was approved by the North-West Multi-Centre Research Ethics Committee and all participants provided written informed consent.

For the MR analysis, we used individual-level imputed data on genetic variants. Genotyping was performed using the Affymetrix UK BiLEVE Axiom array and the Affymetrix UK Biobank Axiom Array.^16,17^ Genotype imputation was based on the Haplotype Reference Consortium and the UK10K haplotype reference panel.^18^

Of the 502,412 UK Biobank participants, we excluded 15,209 individuals because they had missing values on the genetic variants included in our analysis. The remaining 487,203 individuals contributed to the present analysis.

Specific definitions of pre-eclampsia/eclampsia, gestational hypertension, CVD events, and additional variables are outlined in **Supplementary Methods**.

### Statistical analysis

We summarised continuous variables with mean and standard deviation (SD), if normally distributed, and with median and interquartile range, otherwise. Categorical variables were summarised with number and percentage. For all analyses, we used two-sided statistical tests and deemed P-values≤0.05 as statistically significant. Analyses were carried out using R 4.0.5 (The R Foundation, Vienna, Austria).

#### Instrumental variable

For our instrumental variables, we used SNPs for pre-eclampsia/eclampsia and gestational hypertension identified by a recent genome-wide association study (GWAS) by Honigberg et al.^19^ We used effect sizes and standard errors from the discovery analysis, which excluded data from the UK Biobank, allowing two-sample MR.^19^

As illustrated in **Figure S1**, twelve SNPs for pre-eclampsia/eclampsia and seven SNPs for gestational hypertension were identified by the GWAS.^19^ We harmonised the summary-level data to be represented on the same strand. Furthermore, we excluded palindromic SNPs with allele frequencies between 0.45 and 0.55. For both exposures, we excluded one palindromic SNP (rs9855086). To identify related traits, we scanned Phenoscanner^20^ (on February 14^th^ 2023) and extracted all traits associated with the SNPs or any proxies in high linkage disequilibrium (R^2^≥0.8) with P-values≤5×10^−8^ omitting traits identified from the UK Biobank by Neale et al. (http://www.nealelab.is/uk-biobank/). Details on the SNPs included in the present analysis and related traits are provided in **Table S2**.

We measured the strength of our instrumental variable based on the F-statistics of each SNP individually, which ranged from 30 to 58.

#### Primary analysis

We conducted all analyses sex-specific and for women and men combined. For the association between the genetic variants and CVD events, we implemented logistic regression adjusting for age at baseline, sex, if appropriate, and the first 16 genetic principal components, as suggested previously.^21^ We conducted the MR analysis using the R-package *MendelianRandomization*.^22^ In the primary analysis, we applied inverse variance weighted (IVW) regression. We obtained women-to-men ratios of odds ratios (ORs) and 95% confidence intervals (CIs) by subtracting logORs for men from logORs for women and adding sex-specific variances. All ORs in MR analyses are reported per unit increase in the log odds of genetically proxied pre-eclampsia/eclampsia and gestational hypertension.

#### Sensitivity analyses

Sensitivity analyses were conducted similarly to the primary analysis but by applying simple and weighted median regression, MR-Egger, and MR-PRESSO.^23^

Furthermore, we studied associations between genetic variants and CVD events using a Cox proportional hazards model. We used age as underlying timescale and adjusted for sex, if appropriate, and the first 16 genetic principal components. For this sensitivity analysis we excluded all individuals with history of CVD (defined as myocardial infarction or stroke) at baseline. Time-to-event was defined as time to the first CVD event, death, or end of follow-up, whichever occurred first.

Moreover, we conducted a sensitivity analysis, classifying women into ever and never pregnant. We obtained ever-to-never-pregnant ratios of ORs and 95% CIs by subtracting logORs for never pregnant from logORs for ever pregnant women and adding corresponding variances.

#### Secondary analyses

In addition, we conducted a MR analysis using a set of cardiovascular risk factors as outcome variables. We analysed blood pressure-related traits including systolic blood pressure (SBP), diastolic blood pressure (DBP), and age at hypertension diagnosis. Furthermore, we analysed additional CVD risk factors including BMI, total cholesterol, high-density lipoprotein cholesterol, triglycerides, low-density lipoprotein cholesterol, lipoprotein(a), apolipoprotein A1, apolipoprotein B, HbA1c, creatinine, albumin, ALAT, ASAT, GGT, and C-reactive protein. For obtaining genetic associations with risk factors, we used linear regression analysis adjusting for age, sex, if appropriate, and the first 16 genetic principal components,^21^ and divided all cardiovascular risk factors by their SD to enhance the comparison between the individual risk factors. The MR analysis was based on IVW regression.

## Results

### Characteristics of the study population

Baseline characteristics of the included participants are outlined in **Table 1**. The present analysis included 264,160 (54%) women and 223,043 (46%) men. Mean age at baseline was 56.5 (SD 8.1) years. CVD was reported in 13,861 women (7,232 myocardial infarctions; 7,436 strokes) and 27,682 men (19,746 myocardial infarctions; 10,104 strokes).

**Table 1.** Participant characteristics.

| Characteristic | Overall (n=487,203) |  | Women (n=264,160) |  | Men (n=223,043) |  |
| --- | --- | --- | --- | --- | --- | --- |
| | N | Mean ± SD, median [IQR], N (%) | N | Mean ± SD, median [IQR], N (%) | N | Mean ± SD, median [IQR], N (%) |
| Age, years | 487,203 | 56.5 ± 8.1 | 264,160 | 56.4 ± 8.0 | 223,043 | 56.7 ± 8.2 |
| SBP, mmHg | 460,029 | 137.8 ± 18.7 | 249,189 | 135.2 ± 19.2 | 210,840 | 140.9 ± 17.5 |
| DBP, mmHg | 460,034 | 82.2 ± 10.1 | 249,192 | 80.7 ± 10.0 | 210,842 | 84.1 ± 10.0 |
| Hypertension | 474,896 | 224,331 (47.2%) | 264,054 | 117,037 (44.3%) | 210,842 | 107,294 (50.9%) |
| Age at hypertension diagnosis, years | 117,548 | 50.6 ± 9.9 | 54,552 | 50.0 ± 10.8 | 62,996 | 51.2 ± 9.1 |
| Body mass index, kg/m<sup>2</sup> | 485,231 | 27.4 ± 4.8 | 263,213 | 27.1 ± 5.2 | 222,018 | 27.8 ± 4.2 |
| Smoking status | 484,701 |  | 262,851 |  | 221,850 |  |
| Never |  | 265,322 (54.7%) |  | 156,643 (59.6%) |  | 108,679 (49.0%) |
| Ex |  | 168,194 (34.7%) |  | 82,760 (31.5%) |  | 85,434 (38.5%) |
| Current |  | 51,185 (10.6%) |  | 23,448 (8.9%) |  | 27,737 (12.5%) |
| Total cholesterol, mmol/L | 464,313 | 5.7 ± 1.1 | 251,599 | 5.9 ± 1.1 | 212,714 | 5.5 ± 1.1 |
| HDL cholesterol, mmol/L | 424,993 | 1.4 ± 0.4 | 228,604 | 1.6 ± 0.4 | 196,389 | 1.3 ± 0.3 |
| Triglycerides, mmol/L | 463,945 | 1.5 [1.0, 2.1] | 251,460 | 1.3 [1.0, 1.9] | 212,485 | 1.7 [1.2, 2.4] |
| LDL cholesterol, mmol/L | 463,446 | 3.6 ± 0.9 | 251,188 | 3.6 ± 0.9 | 212,258 | 3.5 ± 0.9 |
| Lipoprotein(a), nmol/L | 371,400 | 21.1 [9.6, 61.9] | 201,844 | 22.2 [9.9, 62.0] | 169,556 | 19.8 [9.2, 61.8] |
| Apolipoprotein A1, g/L | 422,655 | 1.5 ± 0.3 | 226,557 | 1.6 ± 0.3 | 196,098 | 1.4 ± 0.2 |
| Apolipoprotein B, g/L | 461,972 | 1.0 ± 0.2 | 250,732 | 1.0 ± 0.2 | 211,240 | 1.0 ± 0.2 |
| HbA1c, mmol/mol | 462,868 | 36.1 ± 6.8 | 250,932 | 35.8 ± 6.0 | 211,936 | 36.5 ± 7.6 |
| Creatinine, μmol/L | 464,085 | 70.4 [61.4, 80.9] | 251,489 | 63.1 [57.0, 70.0] | 212,596 | 80.0 [72.5, 88.4] |
| Albumin, g/L | 425,194 | 45.2 ± 2.6 | 228,687 | 44.9 ± 2.6 | 196,507 | 45.5 ± 2.6 |
| ALAT, U/L | 464,133 | 20.1 [15.4, 27.4] | 251,592 | 17.5 [13.9, 22.9] | 212,541 | 23.9 [18.4, 32.0] |
| ASAT, U/L | 462,594 | 24.4 [21.0, 28.8] | 250,727 | 23.0 [20.0, 26.8] | 211,867 | 26.2 [22.6, 31.0] |
| Gamma-GT, U/L | 464,074 | 26.3 [18.5, 41.0] | 251,496 | 21.4 [16.1, 31.9] | 212,578 | 33.1 [23.7, 50.3] |
| C-reactive protein, mg/L | 463,305 | 1.3 [0.7, 2.8] | 251,162 | 1.4 [0.6, 3.0] | 212,143 | 1.3 [0.7, 2.5] |

Abbreviations: ALAT, alanine aminotransferase; ASAT, aspartate aminotransferase; DBP, diastolic blood pressure; ALAT, alanine aminotransferase; ASAT, aspartate aminotransferase;; HbA1c, glycated hemoglobin; HDL, high-density lipoprotein; IQR, interquartile range; LDL, low-density lipoprotein; SBP, systolic blood pressure; SD, standard deviation.

### MR analysis of genetically proxied HDPs and risk of CVD

#### Primary analysis

As depicted in **Figure 1**, in women and men, respectively, genetically proxied pre-eclampsia/eclampsia was related to a higher risk of CVD with ORs of 1.20 (95% CI 1.01, 1.43) and 1.29 (1.08, 1.53) and to a higher risk of ischaemic CVD with ORs of 1.24 (1.02, 1.52) and 1.32 (1.10, 1.59). We found no significant association with risk of haemorrhagic stroke. Women-to-men ratios of ORs revealed no differences between sexes.

**Figure 1.**
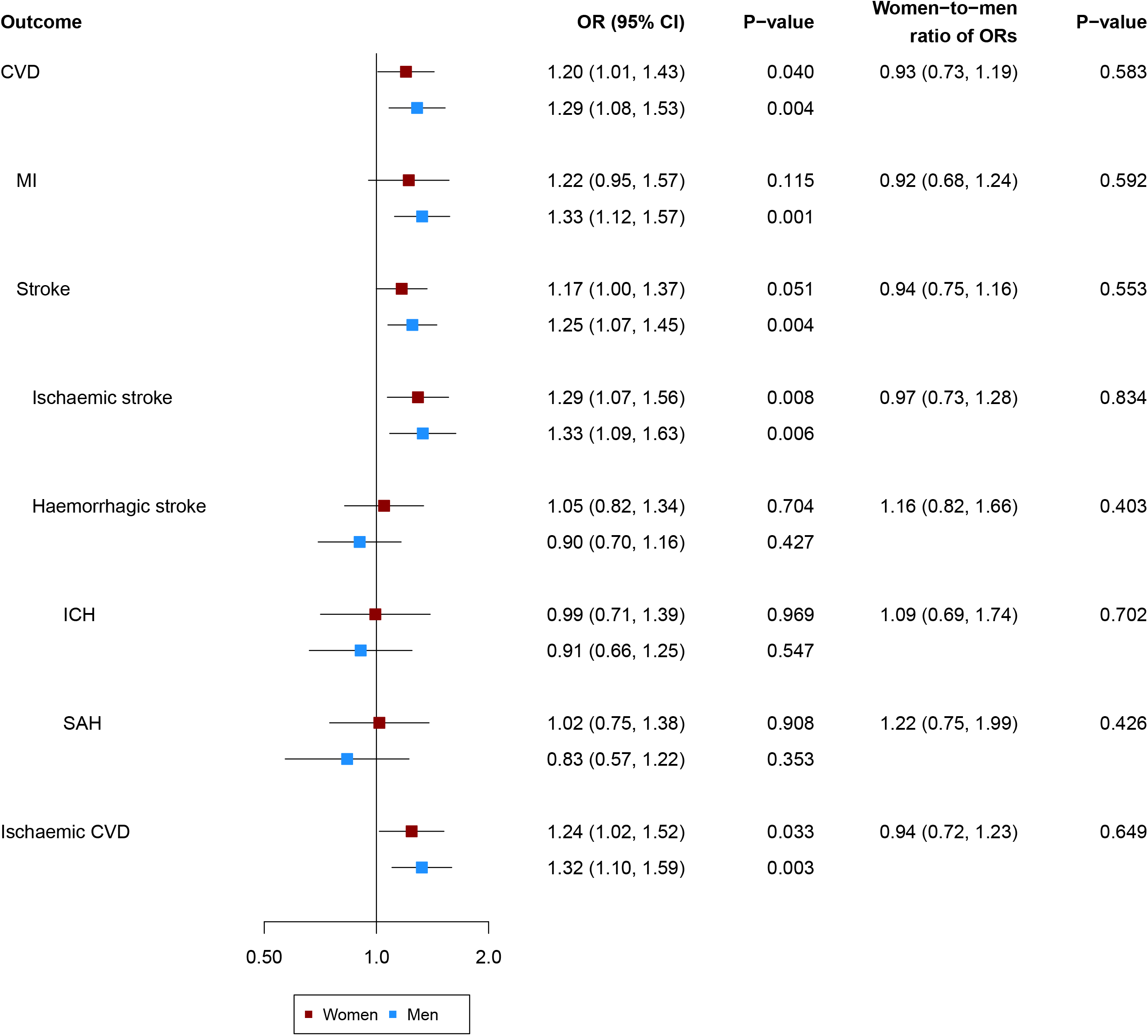
Sex-specific Mendelian Randomisation analysis of genetically proxied pre-eclampsia/eclampsia and risk of cardiovascular events. Results are from inverse variance weighted regression. Models were adjusted for age at baseline and the first 16 genetic principal components. Abbreviations: CI, confidence interval; CVD, cardiovascular disease; ICH, intracerebral haemorrhage; MI, myocardial infarction; OR, odds ratio; SAH, subarachnoid haemorrhage.

**Figure 2** shows the MR analyses for gestational hypertension and CVD events. In women and men, respectively, ORs were 1.22 (1.10, 1.36) and 1.28 (1.16, 1.42) for CVD and 1.22 (1.10, 1.37) and 1.32 (1.19, 1.47) for ischaemic CVD. ORs for haemorrhagic stroke were 1.37 (1.07, 1.77) and 1.12 (0.72, 1.75) in women and men, respectively. Again, women-to-men ratios of ORs were not statistically significantly different from one.

**Figure 2.**
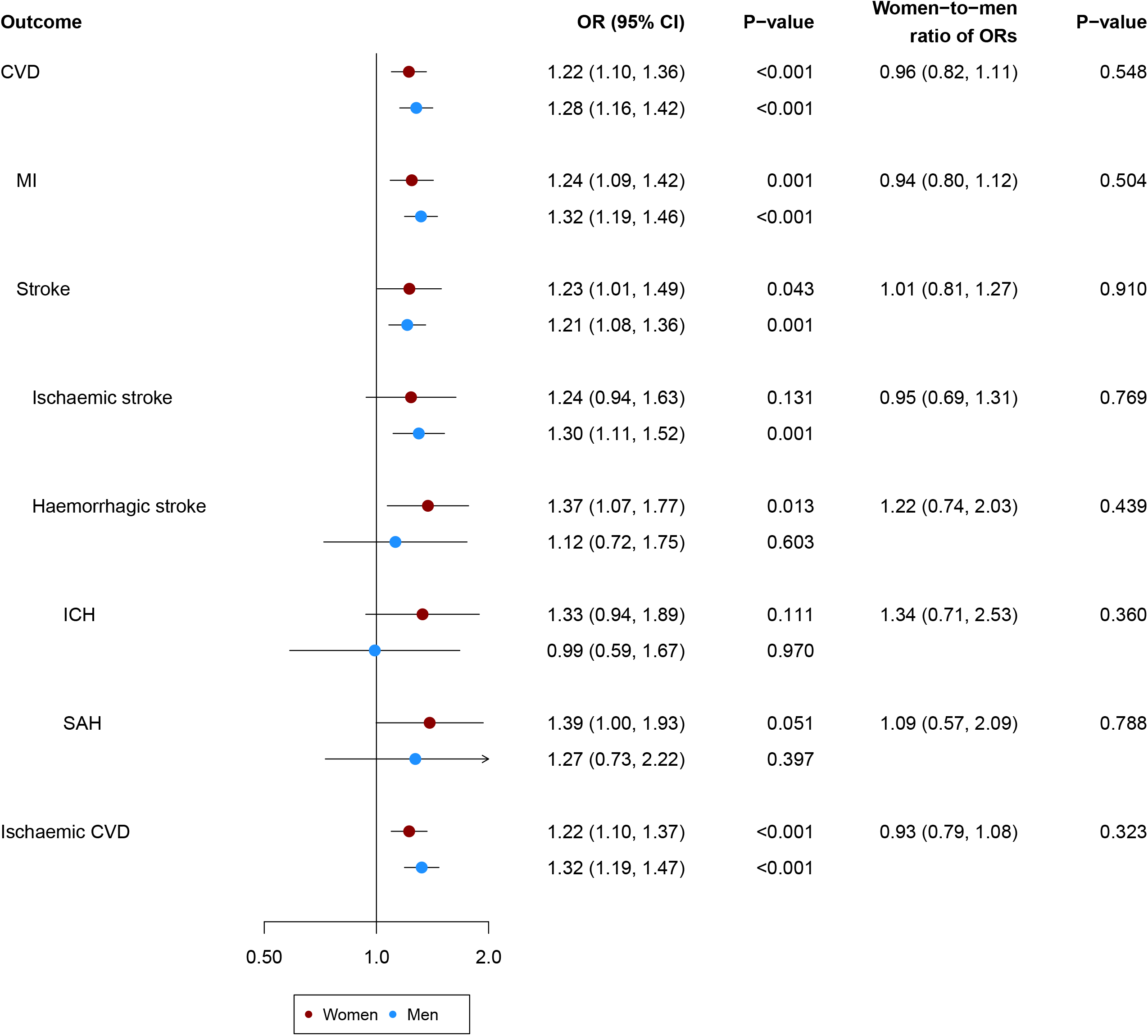
Sex-specific Mendelian Randomisation analysis of genetically proxied gestational hypertension and risk of cardiovascular events. Results are from inverse variance weighted regression. Models were adjusted for age at baseline and the first 16 genetic principal components. Abbreviations: CI, confidence interval; CVD, cardiovascular disease; ICH, intracerebral haemorrhage; MI, myocardial infarction; OR, odds ratio; SAH, subarachnoid haemorrhage.

In sex-combined analyses both genetically proxied HDPs were related to a higher risk of CVD, ischaemic CVD, myocardial infarction, stroke, and ischaemic stroke, but not with haemorrhagic stroke and intracerebral haemorrhage (**Figure S2** and **Figure S3**). Genetically proxied gestational hypertension but not pre-eclampsia/eclampsia was associated with higher risk of subarachnoid haemorrhage.

#### Sensitivity analyses

In sensitivity analyses, applying simple and weighted median regression, results were broadly similar when investigating genetically proxied pre-eclampsia/eclampsia in women (**Figure S4**), men (**Figure S5**), and women and men combined (**Figure S6**). When applying MR-Egger, the OR for intracerebral haemorrhage was 6.21 (1.16, 33.31) in the sex-combined analysis with indication of directional pleiotropy (P-value intercept=0.027). ORs from MR-Egger regression for all other cardiovascular events were not statistically significant. **Table S3** shows the results of MR-PRESSO, which detected outlying variants in the analysis of women, men, and in the sex-combined analysis. The effect sizes remained similar but the association between genetically proxied pre-eclampsia/eclampsia and higher risk of myocardial infarction in women became statistically significant.

When analysing genetically proxied gestational hypertension, results remained robust when implementing simple or median weighted regression for the analysis in women (**Figure S7**), in men (**Figure S8**), and in sex-combined analyses (**Figure S9**). In women, MR-Egger regression yielded a statistically significant intercept (P-value=0.002) when analysing the combined stroke endpoint (**Figure S7**). **Table S3** shows the results of MR-PRESSO, in which genetically proxied gestational hypertension was statistically significantly related to a higher risk of haemorrhagic stroke in the sex-combined analysis (OR 1.32 [1.22, 1.44]).

The Cox regression analysis included 258,416 women and 209,387 men without a history of CVD at baseline. As depicted in **Figure S10**, results remained broadly similar compared to the primary analysis. Genetically proxied pre-eclampsia/eclampsia was related to hazard ratios of 1.13 (0.95, 1.34) and 1.20 (1.01, 1.44) for CVD in women and men, respectively, with no significant differences between the sexes. Hazard ratios for CVD were 1.17 (1.03, 1.32) in women and 1.25 (1.13, 1.38) in men, when analysing genetically proxied gestational hypertension (**Figure S11**). Results of the sex-combined analysis are provided in **Figure S12** and **Figure S13** and were also comparable to the findings of the primary analysis.

**Figure S14** and **Figure S15** show the MR analyses by history of pregnancy. The excess risk for ischaemic stroke for genetic liability to gestational hypertension was higher in ever compared to never pregnant women with a ratio of ORs of 1.84 (1.04, 3.24). We found no significant differences when analysing pre-eclampsia/eclampsia and gestational hypertension and other CVD events.

### MR analysis of genetically proxied HDPs with blood pressure-traits and cardiovascular risk factors

**Figure 3** shows the associations between genetically proxied pre-eclampsia/eclampsia and a range of cardiovascular risk factors. Genetically proxied pre-eclampsia/eclampsia was related to higher levels of SBP and DBP and younger age at hypertension diagnosis. Furthermore, we found a significant association with lower levels of total cholesterol in both women and men. There were no sex-differences in any of the associations.

**Figure 3.**
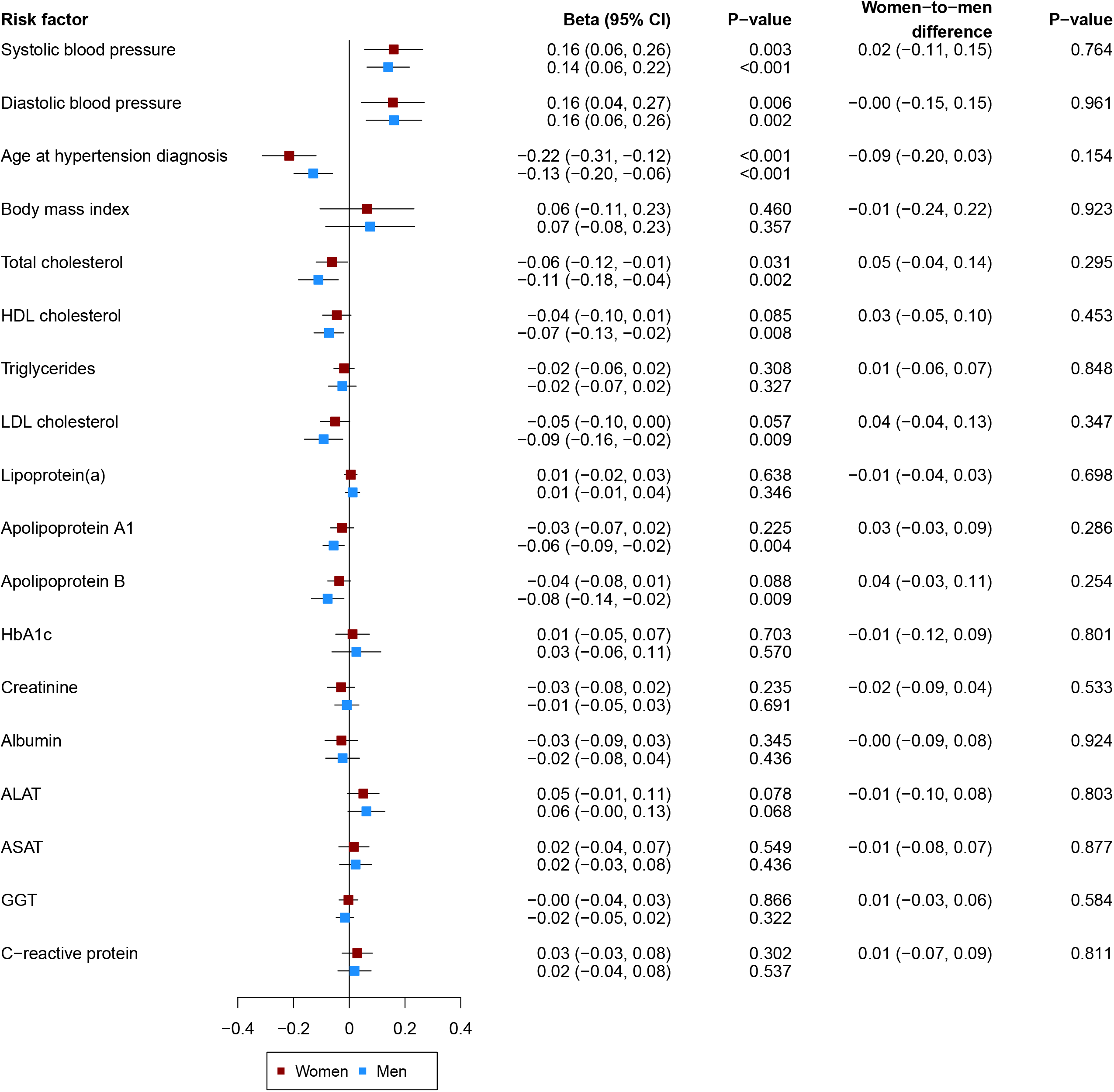
Sex-specific Mendelian Randomisation analysis of genetically proxied pre-eclampsia/eclampsia and cardiovascular risk factors. Results are from inverse variance weighted regression Results are from inverse variance weighted regression. Models were adjusted for age at baseline and the first 16 genetic principal components. Abbreviations: ALAT, alanine aminotransferase; ASAT, aspartate aminotransferase; CI, confidence interval; GGT, gamma glutamyltransferase; HbA1c, glycated haemoglobin; HDL, high-density lipoprotein; LDL, low-density lipoprotein.

Relations between genetically proxied gestational hypertension and cardiovascular risk factors are depicted in **Figure 4**. Genetically proxied gestational hypertension was associated with higher levels of SBP and DBP and younger age at hypertension diagnosis in women and men with no statistically significant differences between women and men.

**Figure 4.**
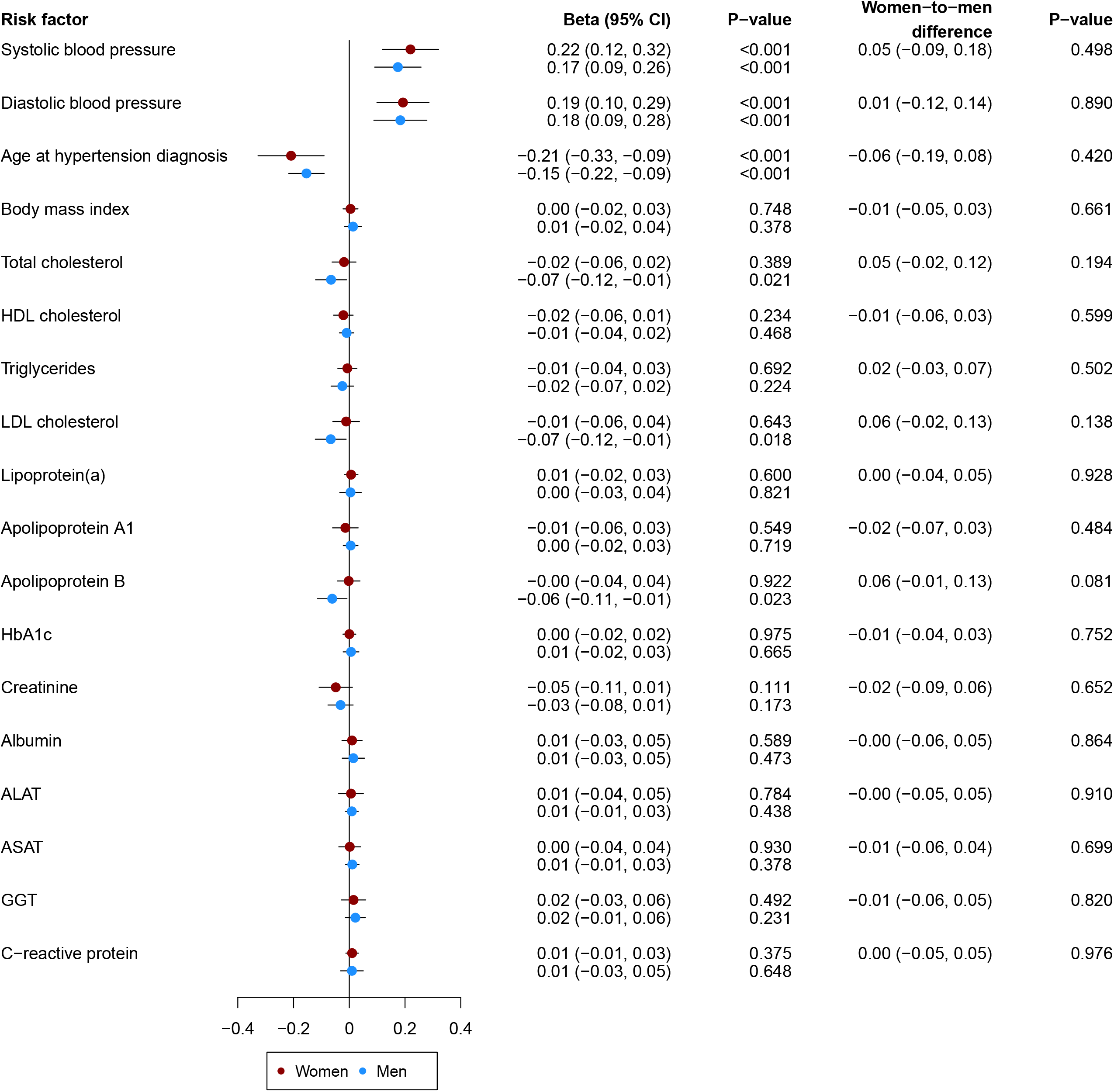
Sex-specific Mendelian Randomisation analysis of genetically proxied gestational hypertension and cardiovascular risk factors. Results are from inverse variance weighted regression. Models were adjusted for age at baseline and the first 16 genetic principal components. Abbreviations: ALAT, alanine aminotransferase; ASAT, aspartate aminotransferase; CI, confidence interval; GGT, gamma glutamyltransferase; HbA1c, glycated hemoglobin; HDL, high-density lipoprotein; LDL, low-density lipoprotein.

Sex-combined analyses for genetically proxied pre-eclampsia/eclampsia and gestational hypertension are shown in **Figure S16** and **Figure S17**, respectively. Again, both genetically proxied pre-eclampsia/eclampsia and gestational hypertension were related to higher levels of SBP and DBP and younger age at hypertension diagnosis. Additionally, genetically proxied pre-eclampsia/eclampsia was associated with lower levels of several lipid parameters.

## Discussion

In this analysis, we found associations between genetic liability to pre-eclampsia/eclampsia and gestational hypertension and higher risk of CVD in both women and men. Furthermore, both HDPs were related to higher levels of SBP and DBP and younger age at hypertension diagnosis.

### Findings from previous studies

A positive association between HDPs and risk of CVD has previously been reported by observational studies. A large-scale meta-analysis showed that moderate and severe pre-eclampsia are associated with a higher risk of maternal CVD, with ORs of 2.24 (1.72, 2.93) and 2.74 (2.48, 3.04), respectively.^24^ The same is the case for gestational hypertension, for which an OR for maternal CVD of 1.67 (1.28, 2.19) has been reported.^24^ Moreover, a phenome-wide association analysis showed strong associations between polygenic risk scores of HDPs with CVD risk factors (such as hypertension, diabetes mellitus, hyperlipidaemia, and obesity) and CVD events (including ischaemic heart disease and coronary atherosclerosis) in women and men.^19^ In addition, a recent MR analysis, which only included women for genetic associations with HDPs, showed genetically proxied HDPs to be related to a higher risk of coronary artery disease or ischaemic stroke.^11^ Furthermore, genetically proxied pre-eclampsia and gestational hypertension were associated with a higher risk of coronary artery disease but not with risk of ischaemic stroke.^11^ This is not in line with the results of the present study, in which we did find a significantly higher risk for ischaemic stroke for genetically proxied pre-eclampsia/eclampsia in women. However, it has to be noted that the previous MR study only included data from one database for obtaining genetic associations with genetically proxied HDPs and used sex-combined effect sizes for the genetic associations with CVD events, which could potentially explain these differences.^11^ In the present MR study, we (1) used effect sizes for the genetic association with the exposure from a large-scale GWAS that meta-analysed results of multiple studies, (2) analysed sex-specific effect sizes for the genetic association with the outcome, (3) included analyses restricted to men, and (4) studied associations with blood pressure-, lipid-, liver-, and kidney-related traits.

### HDPs and blood pressure

Per definition, hypertension is the major component of HDPs. In the present analysis, we showed that genetically proxied pre-eclampsia/eclampsia and gestational hypertension are significantly associated with both higher SBP and DBP later in life. Genetic correlation between HDPs and blood pressure has previously been demonstrated.^19^ Notably, the genetic correlation between gestational hypertension and SBP was even higher than between SBP and DBP.^19^ A previous MR analysis showed that higher genetically predicted SBP was also related to an elevated risk for pre-eclampsia/eclampsia.^25^ Moreover, higher levels of genetically proxied blood pressure have been related to an increased risk of CVD in both women and men.^26^ In case blood pressure lies on the causal pathway between pre-eclampsia/eclampsia and gestational hypertension and CVD, we speak of vertical pleiotropy, which does not bias the findings of our MR analysis. However, we cannot fully exclude horizontal pleiotropy, which would violate the assumptions of MR. When applying MR-Egger, we found no significant intercepts in the relation of pre-eclampsia/eclampsia and CVD, which indicates absence of directional pleiotropy. We did find a significant MR-Egger intercept in the association between genetically proxied gestational hypertension and risk of stroke and ischaemic stroke in women. However, it has previously been discussed that MR-Egger results can be influenced by outlying variants.^27^ When we further studied the relation between genetically proxied gestational hypertension and risk of stroke and ischaemic stroke applying MR-PRESSO, the findings of our primary analysis remained robust.

### Differences between ischaemic and haemorrhagic events

Our MR analysis revealed both pre-eclampsia/eclampsia and gestational hypertension to be related to ischaemic CVD. Contrarily, we found no statistically significant association between pre-eclampsia/eclampsia and risk of haemorrhagic stroke. These findings are not in line with results from previous observational studies. In a US database it has been shown that pre-eclampsia/eclampsia was associated with an OR for pregnancy-related intracerebral haemorrhage of 10.39 (8.32, 12.98).^28^ Moreover, a study from Taiwan reported a hazard ratio for haemorrhagic stroke of 3.50 (2.31, 5.29) related to pre-eclampsia.^29^ Another analysis in Canadian women found pre-eclampsia to be related to a higher risk of haemorrhagic stroke.^30^ Therefore, our findings suggest that the results of observational studies may be biased by potential confounding factors affecting the relation between pre-eclampsia/eclampsia and risk of haemorrhagic stroke. However, especially our sex-specific analysis may have limited statistical power to analyse haemorrhagic stroke events, which could have prevented us from finding statistically significant associations. Nevertheless, in sex-combined analyses, we also did not find significant association of genetically proxied pre-eclampsia/eclampsia and occurrence of haemorrhagic stroke. In contrast, we did find a statistically significant association between genetically proxied gestational hypertension and the risk of subarachnoid haemorrhage in our sex-combined analysis. This indicates potential differences between the roles of pre-eclampsia/eclampsia and gestational hypertension in the risk for haemorrhagic stroke.

### HDPs in men and nulligravidae

Our MR analysis also included men and nulligravidae, who can have the genetic liability to HDPs although the underlying phenotype can never express. Nevertheless, we found genetic liability to pre-eclampsia/eclampsia and gestational hypertension to be related to a higher risk of CVD events in men. Robillard et al. previously suggested the importance paternity in the development of pre-eclampsia and reported a significantly higher risk for pre-eclampsia in new paternity multiparas compared to same paternity multiparas and primiparas.^31^ To further investigate how the genetic liability to HDPs expresses in men, we studied their relationship with a range of cardiovascular risk factors. Interestingly, genetically proxied HDPs were also related to younger age at hypertension diagnosis in men. A recently conducted phenome-wide association study investigated the relation between polygenic risk scores for pre-eclampsia/eclampsia and gestational hypertension with >1,000 phenotypes in men and also reported hypertension to be one of the key associated characteristics.^19^ This is indicative that earlier hypertension is a main driver of the underlying mechanisms leading to a higher CVD risk in males. We also analysed the genetic liability to HDPs and CVD in nulligravidae. Relations were similar in never and ever pregnant women for all CVD events, but ischaemic stroke, as we found a statistically significantly higher excess risk for ischaemic stroke related to the genetic liability to gestational hypertension for ever compared to never pregnant women. However, it should be noted that statistical power for the analysis of nulligravid women was limited. Consequently, the role of genetic liability to HDPs in CVD risk in nulligravidae needs further investigation.

### Strengths and limitations

Our study has several strengths. We used data from UK Biobank, which provided adequate statistical power to conduct the MR analyses. Furthermore, we built our genetic instruments on a comprehensive large-scale GWAS that meta-analysed data from multiple studies.^19^ Moreover, due to the use of individual-participant data from the UK Biobank, we were able to study sex-specific associations of genetic variants with CVD events. In addition, we conducted several sensitivity analyses including different methods of MR and a Cox-regression analysis based on time-to-event data. Our analysis also has limitations. First, the effect sizes for the genetic association with the exposure variable were based on individuals of different ancestries, while the UK Biobank majorly includes individuals of European ancestry. Furthermore, we did not have adequate statistical power for analysing nulligravid women, limiting the interpretation of our findings. Moreover, MR analysis relies on three assumptions. The first assumption is that the instrumental variable is associated with the exposure. Phenotypic data on pre-eclampsia/eclampsia and gestational hypertension were not available in our dataset. Consequently, we were not able to investigate whether the genetic instruments correlated with the phenotype in our study. However, we calculated F-statistics for each SNP individually and all of them were ≥30 indicating that we have sufficiently strong instruments. In addition, these SNPs were obtained from an independent GWAS, which replicated the majority of associations in additional cohorts, supporting our assumption of having robust associations with the exposures. The second assumption is that the genetic instrument is not associated to any confounding factor. We studied the association of genetically proxied pre-eclampsia/eclampsia and gestational hypertension with several cardiovascular risk factors. We found significant associations between genetically predicted HDPs and lower levels of lipid parameters. As lower levels of some lipid parameters (e.g., total cholesterol) are known to be related to a lower cardiovascular risk,^32^ while others have been related to a higher cardiovascular risk (e.g., high-density lipoprotein cholesterol),^33^ the specific role of lipid levels on the pathway between HDPs and risk of cardiovascular events needs to be further investigated. The third assumption is that the genetic instrument can influence the outcome only via the exposure. To study this assumption, we conducted MR-Egger regression and MR-PRESSO, which indicated potential directional pleiotropy for single CVD events as discussed in detail above.

## Conclusion

Genetic liability to HDPs including pre-eclampsia/eclampsia and gestational hypertension is associated with higher CVD risk, implying biological mechanisms relating to these disorders are causally related to CVD risk in both women and men.

## Perspectives

This MR study demonstrated that genetic liability to pre-eclampsia/eclampsia and gestational hypertension is related to a higher risk for CVD. Additionally, genetic liability to HDPs was related to higher blood pressure levels and younger age at hypertension diagnosis. The findings were similar in women and men. This implies that biological mechanisms related to HDPs are causally related to CVD risk. Furthermore, it suggests that both HDPs and CVD events may be expressions of the same disease pathway at different stages in life and that HDPs may be an indicator for individuals at higher CVD risk later in life. While pregnancy allows identification of women at increased risk for future CVD, men and nulligravidae with the same genetic liability to HDPs may not have the opportunity to present at an earlier stage in life.

## Data Availability

Summary-level data from the GWAS have been published previously (doi: 10.1101/2022.11.30.22282929). Data from the UK Biobank can be requested via their website (https://www.ukbiobank.ac.uk/enable-your-research/apply-for-access).

## Acknowledgements

This research has been conducted using the UK Biobank Resource (Application Number 29916). We are extremely grateful to all participants in the UK Biobank.

## Data availability

Summary-level data from the GWAS have been published previously.^19^ Data from the UK Biobank can be requested via their website (https://www.ukbiobank.ac.uk/enable-your-research/apply-for-access).

## Novelty and Relevance

### What is New?

- This Mendelian Randomisation study investigated the relation between genetically predicted hypertensive disorders of pregnancy and the risk of developing cardiovascular disease and several cardiovascular risk factors.

### What is Relevant?

- The study found genetic liability to pre-eclampsia/eclampsia and gestational hypertension to be linked to a higher risk of cardiovascular disease events in both women and men.
- Earlier hypertension appears to be a main driver behind the relation of hypertensive disorders of pregnancy and increased cardiovascular risk.

### Clinical/Pathophysiological implications?

- Hypertensive disorders of pregnancy seem to be an indicator for individuals at higher cardiovascular disease risk later in life.

